# A Modular Architecture for Dynamic Evaluation of Best-Practice Care

**DOI:** 10.64898/2026.10.03.26364324

**Authors:** Minh Trang Hoang, Tim Shaw, Christina Igasto, Candice Donnelly, Amith Shetty, Malcolm Pradhan

## Abstract

**Objective:** Retrospective evaluation of clinical pathway adherence using electronic medical record (eMR) data is limited by heterogeneous data structures and statistical measures that do not account for patient-specific context, care sequence, or timing. We designed a modular event-based architecture to dynamically evaluate the New South Wales Adult Sepsis Pathway over time.

**Method:** Data from eMRs were collected from adult emergency department encounters across four hospitals between July 2022 and June 2024 and processed through three sequential layers: event-stream transformation, pathway-relevance filtering, and state-based pathway evaluation. Heterogeneous records were standardized and organized into a chronological timeline for each encounter. The framework then updated the pathway’s state as new clinical information became available and assessed whether recommended care was delivered within the relevant timeframe and sequence.

**Result:** The architecture processed 451,124 encounters. This included 57,631,084 clinical events extracted and transformed into encounter-level event streams. After filtering for pathway relevance, 43,951,146 events remained (76.3%). A total of 129,907 (34.6%) encounters entered the sepsis pathway, 50,118 (38.6%) entered YELLOW state, indicating progression to suspected sepsis, and 3,349 (2.6%) encounters progressed to RED state from GREEN, indicating the initial shock. Of 50,118 encounters entering YELLOW state, 2,426 (4.8%) progressed to shock, 8,700 (17.4%) encounters were recognized with sepsis through a blood culture order, and 4,287 (8.6%) encounters were recognized with sepsis through an antibiotic order. This demonstrates the architecture’s ability to prioritize higher-acuity states during evaluation rather than going through an intermediate state.

**Conclusion:** The architecture developed shows the feasibility of transforming fragmented routine eMR data into a reusable longitudinal representation that supports context-dependent pathway evaluation. Separating data preparation from pathway rules improves transparency, maintainability, and adaptability and provides a foundation for future interoperable, near-real-time pathway monitoring

## Introduction

Clinical practice guidelines (CPGs) are essential in promoting evidence-based care, increasing hospital efficiency, decreasing operational costs, and improving patient outcomes [1-4]. Clinical pathways (CPs) translate CPGs’ recommendations into an organized sequence of time- and condition-dependent care processes that can be followed and evaluated over time [5, 6]. Existing literature has shown that implementation of CPs improves patient care quality, identifies practice variations, and supports care coordination and delivery [5, 7, 8]. Ultimately, evaluating whether recommended care is delivered in accordance with CPs indicates healthcare quality and reflects the healthcare system’s efforts to implement evidence-based care systematically.

Recently, there has been increased interest in using electronic medical record (eMR) data to evaluate adherence to best practices, owing to its potential to provide large volumes of longitudinal, structured clinical data [9-11]. However, despite advances in data infrastructure and analytics, several challenges limit the effectiveness of using eMR data to evaluate adherence to best-practice care [12]. Because eMR systems are primarily designed to support care delivery, documentation, and administration rather than pathway evaluation, clinically important variables for pathway evaluation such as disease onset, decision points, and reasons for treatment may not be explicitly represented [13, 14]. Data may also be incomplete, inconsistently structured, recorded retrospectively, and distributed across multiple source systems [15-17]. These challenges show that the current representation of eMR data is limited in its ability to capture the nuanced temporal complexity of clinical processes as patients’ conditions evolve.

Traditional health services research relies on discrete indicators to assess whether recommended care was delivered within a set period [18-20]. While useful for reporting and benchmarking, these measures often offer a static view and may not capture the clinical reasoning behind differences between recommended and actual care. Computational methods, such as rule-based systems, computer-interpretable guidelines, process models, and conformance checking have been developed to evaluate guideline adherence [21-24]. However, their implementation in clinical practice remains challenging because of limitations in usability, integration, and interoperability across healthcare systems [25]. Existing adherence approaches also less commonly represent adherence as an evolving process in which changes in the patient’s clinical condition alter which care recommendations are applicable and how adherence should be evaluated. Consequently, there is a gap in understanding how care recommendations in CPs are followed and adapted to a patient’s evolving condition, rather than analyzing each event in isolation or applying uniform rules to all patients.

Sepsis provides a clinically relevant example of the challenges involved in evaluating care that must adapt to a patient’s evolving condition over time. Prompt management to prevent deterioration reduces mortality, length of stay, and healthcare costs [26-28]. In New South Wales (NSW), Australia, the SEPSIS KILLS program offers a standardized clinical pathway, the Adult Sepsis Pathway (ASP), aiding clinicians to recognize and respond to patients at risk of developing sepsis [29]. The ASP details the signs and symptoms of sepsis, such as suspected infection, suspected sepsis, or septic shock, and outlines care recommendations and their timeframes for adult patients. As a result, the ASP reflects a dynamic process in which applicable recommendations and care priorities change with the patient’s condition, and not all recommendations in the pathway apply to every patient. Evaluating adherence dynamically may therefore provide a more context-sensitive representation of care than applying the pathway as a uniform binary checklist.

A generalizable approach to dynamic guideline adherence evaluation requires two linked capabilities. First, heterogeneous eMR data must be transformed into a common longitudinal representation that preserves the timing, sequence, and clinical meaning of events. Second, the evaluation approach must maintain and update a representation of the patient’s clinical condition and their position within the pathway as care unfolds. Patient state represents the patient’s evolving clinical condition based on recorded observations and other relevant clinical events. Pathway state represents the patient’s current position within the clinical pathway, including which care recommendations have been activated, fulfilled, remain outstanding, or are no longer applicable, together with their required timeframes. Maintaining state enables the evaluation to account for previous events when determining which care recommendations are applicable as the encounter progresses, rather than evaluating each event independently or applying all recommendations as a static checklist.

In this study, we designed and demonstrated a modular architecture for dynamically evaluating care delivered under the ASP using routinely collected eMR data. The specific objectives were:

- Transform heterogeneous tabular eMR data into standardized encounter-level streams of clinically meaningful events that reconstruct the sequence and timing of care and enable comparison with the care recommendations specified in the ASP.
- Use these event streams to evaluate adherence by tracking changes in patient and pathway states, identifying where specific care recommendations became relevant, and determining whether the expected care was delivered in the appropriate order and within the specified timeframe.

## Method

This study was approved by the Western Sydney Local Health District Human Research Ethics Committee (2019/ETH09607).

### Study design, setting and data source

We conducted a retrospective systems design and development study of a modular computational architecture for evaluating care delivered under the ASP using routinely collected eMR data. The study comprised two linked components: transforming heterogeneous eMR tables into standardized encounter-level event streams, followed by a time-based, state-based evaluation of pathway progression and care delivery.

We extracted and analyzed eMR data from four hospitals within the Western Sydney Local Health District (WSLHD), Australia. The district serves a multicultural population of over a million residents. Each patient presentation to the Emergency Department (ED) of any hospital within WSLHD on a separate occasion during the study period was classified as an encounter. Eligible encounters included all presentations of patients aged 16 years and older between July 2022 and June 2024. Encounters with documented pregnancy were excluded. Source data included encounter visit records, triage assessment, vital-sign observations, laboratory orders and results, medication orders and administrations, and structured clinical forms. Data were cleansed and de-identified before provision to the analysis team. Records were linked using hashed patient and encounter identifiers. Available timestamps were used to reconstruct the order and timing of clinical events within each encounter.

### Clinical pathway specification

The ASP [29] was translated into a computable specification comprising clinical concepts, patient state variables, pathway states, transition rules, and state-specific care recommendations. Clinical concepts included prior risk factors, infection-related presenting problems, physiological observations, investigations, and medication events. These concepts were defined independently of how they appeared in source tables and subsequently linked to local eMR terms through semantic mapping.

The architecture maintained two related forms of state. Patient state represented the clinical information available about the patient encounter at the review point, whereas pathway state represented the patient’s current position within the ASP and retained memory of previous pathway transitions. The pathway states and their operational triggers are summarized in Table 1. State transitions were determined from combinations of physiological abnormalities, infection-related signals, and clinical actions based on the Systemic Inflammatory Response Syndrome (SIRS) criteria as outlined in the ASP [29]. Where pathway criteria depended on information not explicitly documented in structured data, such as clinicians’ suspicion of sepsis, observable structured-data proxies were used. These operational definitions were intended as computational approximations of pathway logic rather than direct measures of clinical reasoning. State transitions were time-stamped and irreversible once a higher-acuity state was reached, thereby preserving care recommendations activated by earlier deterioration.

**Table 1.** Clinical interpretation and operationalization of pathway states.

| State | Clinical interpretation | Operational trigger |
| --- | --- | --- |
| START | Encounter begins before pathway assessment | First time-stamped encounter event |
| GREEN | Suspected infection with sufficient concern to enter sepsis surveillance | Combination of infection-related presenting problems, prior risk factors, and any type of cultures ordered |
| YELLOW | Physiological deterioration suggests suspected sepsis | Moderate physiological abnormality pattern meeting YELLOW criteria in the ASP |
| RED | Severe deterioration suggests confirmed sepsis or septic shock, requiring urgent sepsis management | Severe physiological abnormality escalation pattern meeting RED criteria in the ASP |
| EXIT | Pathway evaluation ends | First antibiotic administration or encounter completion |

Pathway state determined which care recommendations were applicable, when their completion windows commenced, and when evaluation ended. For example, when a patient was suspected of sepsis or septic shock, time-critical recommendations such as antibiotic administration, lactate measurement, laboratory investigations, blood culture order, IV fluid administration, and regular vital signs monitoring were activated. Each recommendation was specified by its qualifying evidence, completion window, temporal dependencies, and closure rule. When a recommendation was activated, the engine could examine relevant preceding and subsequent events to ensure that clinically appropriate care delivered before computable escalation could be recognized.

### Computational architecture

The architecture was implemented as a retrospective monthly batch-processing pipeline with three sequential layers: (1) Event-stream transformation, (2) Pathway-relevant event filtering, and (3) Evaluation framework (Figure 1, Panel A). Each layer operated independently on the output of the preceding layer. The architecture was developed for retrospective evaluation. No real-time data ingestion or clinical decision support functionality was deployed

**Figure 1.**
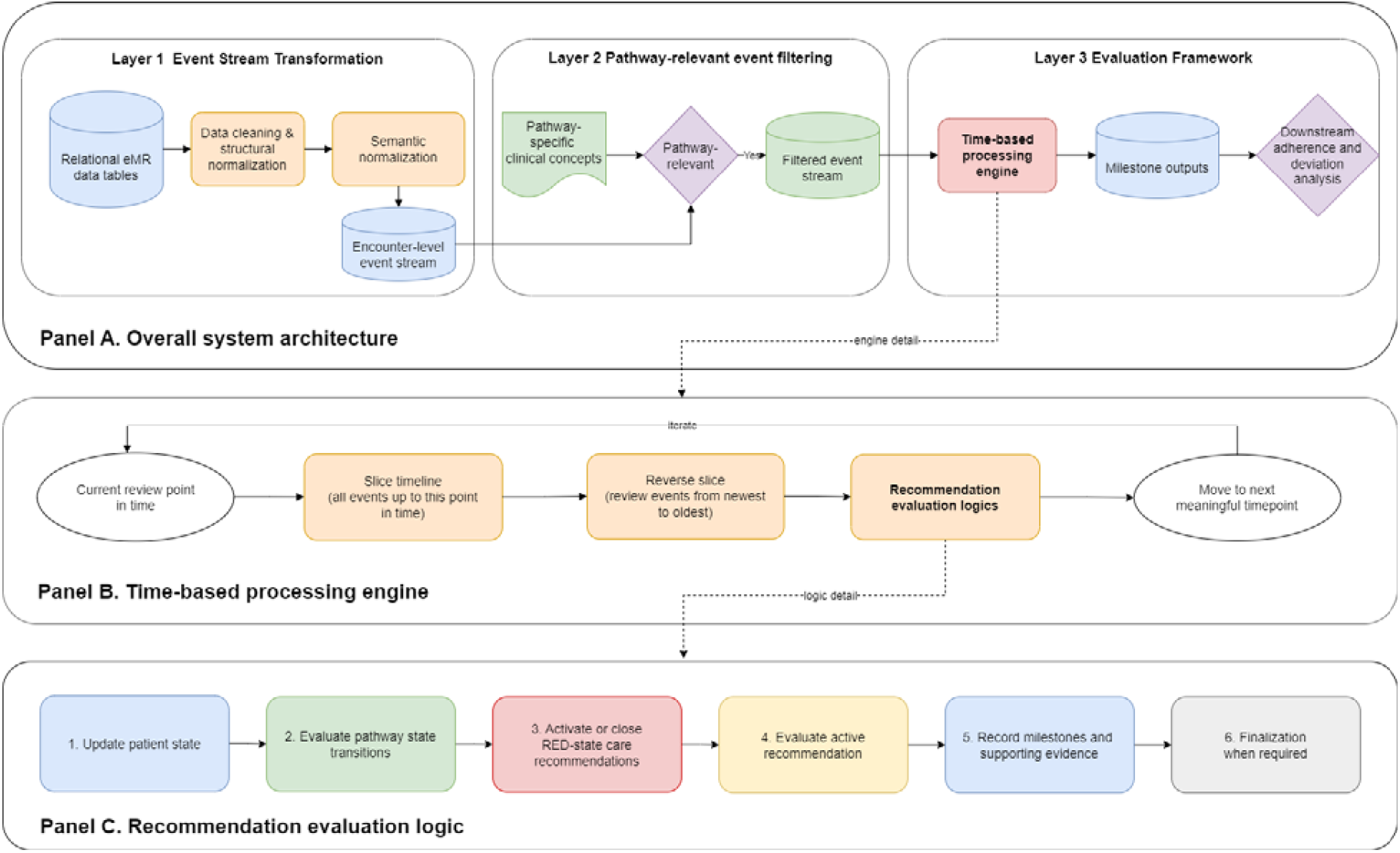
Overall system architecture. Panel A describes the overall system architecture, Panel B depicts the zoom-in of the time-based processing engine, and Panel C illustrates the care recommendation-evaluation logic executed at each review point.

### Event stream transformation

As eMR data tables differed in structure and terminology, they could not be directly processed by the evaluation framework. Therefore, they were converted into a standardized longitudinal event representation through structural normalization, semantic normalization, and chronological assembly. Structural normalization mapped each record to standard fields describing event type, patient and encounter identifiers, event identifier, timestamp, code, value, unit, and pathway relevance (Table 2). Timestamps were converted to a common format. Exact duplicates were removed using encounter identifier, event identifier, timestamp, and value, while repeated measurements at different times were retained.

**Table 2.** Standardized fields used to represent individual events within encounter-level event streams.

| Field | Description | Example |
| --- | --- | --- |
| resourceType | Type of clinical event | Patient, Encounter, Observation, |
| subject | The hashed unique patient identifier. Used to identify multiple episodes of care for a patient if available |  |
| encounter | The hashed unique encounter identifier used to link multiple clinical events within an encounter |  |
| id | Unique event identifier, enabling linkage between related events such as pathology orders and results, or medication requests and administrations | Medication order ID, laboratory order ID |
| effectiveDateTime | Time stamp of clinical event | Time of antibiotic order, time of antibiotic administration |
| code | The name describing the event | Triage category, systolic blood pressure, Full blood count |
| value | Numeric values for clinical observations and pathology results, categorical strings for pre-defined presenting problem, ordered status, medication names | 150, Female |
| valueUnit | measurement unit corresponding to numeric values in value field | mmHg, years |
| pathwayRelevant | Indicator denoting whether the data object was relevant to the sepsis clinical pathway, based on the list of extracted clinical variables |  |

Semantic normalization mapped heterogeneous source terms to consistent clinical concepts. For example, locally recorded IV fluid products were mapped to a common IV fluid concept. Clinical mappings were reviewed by clinicians for local validity. Vital signs documented within a 3-minute interval were grouped as one observation set. Alert, Voice, Pain, Unresponsive (AVPU) values were converted to Glasgow Coma Scale (GCS) score using the method described by Ramgopal S. et al. [30]. Medication orders and medication administrations were linked using the corresponding event identifiers where possible.

Normalized events were grouped by encounter and ordered by their clinically relevant timestamp to form an encounter-level event stream. No unrecorded clinical actions were imputed; absence of an event was interpreted as absence of documented evidence. This representation preserved the temporal sequence and the clinical meaning of events while harmonizing heterogeneous records from different clinical domains

### Pathway-relevant event filtering

Event streams were filtered to retain concepts required by the computable ASP specification as outlined above, including physiological observations, infection-related presenting problems, laboratory investigations, blood culture, diagnoses, antibiotic and IV fluid events. This layer reduced nonessential transactional data while preserving a reusable general event-stream structure for pathway evaluation

### Evaluation framework

Within the framework, each filtered event stream was processed chronologically using a time-based processing engine (Figure 1, Panel B). A temporal pointer was initialized at encounter start and advanced in short intervals to accommodate near-synchronous documentation At each review point, all events were examined with the most recent events first so that the current patient status could be inferred from the latest available information. The care recommendation-evaluation logic was then applied to the latest available information as shown below

#### Care recommendation-evaluation logic

At each review point, the processing engine applied a predefined sequence of operations to update the patient state, determine the pathway state, and evaluate the care recommendations that were applicable at that time (Figure 1, Panel C). The sequence comprised six steps: updating patient state, evaluating pathway state transition, activating or closing state-specific care recommendations, evaluating active obligations, recording milestone outcomes and supporting evidence, and finalizing obligations when no further evaluation was required.

1. Update patient state Patient state was dynamically reassessed using available clinical data for risk assessment at each review point. Risk assessment integrated physiological indicators derived from vital signs and laboratory results, as well as contextual indicators such as suspected infections and prior risk factors, as specified in the ASP. Variations in risk status were documented and employed to guide state transition.
2. Evaluate pathway state transitions The updated patient state was then evaluated against the pathway transition rules. Transition rules were assessed according to clinical priority, with criteria for the highest-acuity state evaluated before criteria for lower-acuity states. This allowed the engine to move a patient directly to a higher-acuity state when severe evidence was present, without requiring sequential progression through every intermediate state. For each transition, the engine recorded the time of transition, the previous and new pathway states, and the clinical evidence responsible for the change
3. Activating and closing RED-state care recommendations Following pathway state evaluation, the engine activated or closed the care recommendations associated with the current state. An activated recommendation record included the activation time, expected clinical action, completion window, and any temporal dependencies.
4. Evaluate active care recommendations At each review point, each active care recommendation was evaluated using its own completion criteria. The completion status of a recommendation could be pending if its completion window was open and no qualifying evidence had yet been identified. A recommendation was recorded satisfied when the required evidence was present within the applicable temporal and clinical conditions. Repeated recommendations like regular vital signs monitoring could also be recorded as partially completed when only some of the expected evidence was available. Once the allowable completion period elapsed without sufficient qualifying evidence, the care recommendation was classified as missed according to its predefined evaluation rule.
5. Record milestones and supporting evidence The engine generated a milestone record whenever a care recommendation was completed or finalized. Each milestone captured timing, final evaluation status, and supporting evidence. Retaining supporting evidence provides an auditable link between each adherence determination and the underlying eMR events, rather than producing an outcome without explaining how it was derived.
6. Finalization Finalization occurred when the treatment defining the end of the evaluation period was administered, or the encounter was completed. At finalization, unresolved recommendations were assigned to their final status, and all relevant supporting evidence was retained. The resulting milestone-level records served as the structured outputs used for downstream adherence and deviation analysis.

## Results

### Cohort characteristics and data summary

Between July 2022 and June 2024, 451,124 encounters were extracted from the eMR system. We excluded 1,824 (0.47%) encounters with confirmed pregnancy status and 1,731 (0.44%) without valid encounter timestamps. After excluding encounters with invalid age, 388,047 (86.0%) encounters were included for evaluation (Figure 2).

**Figure 2.**
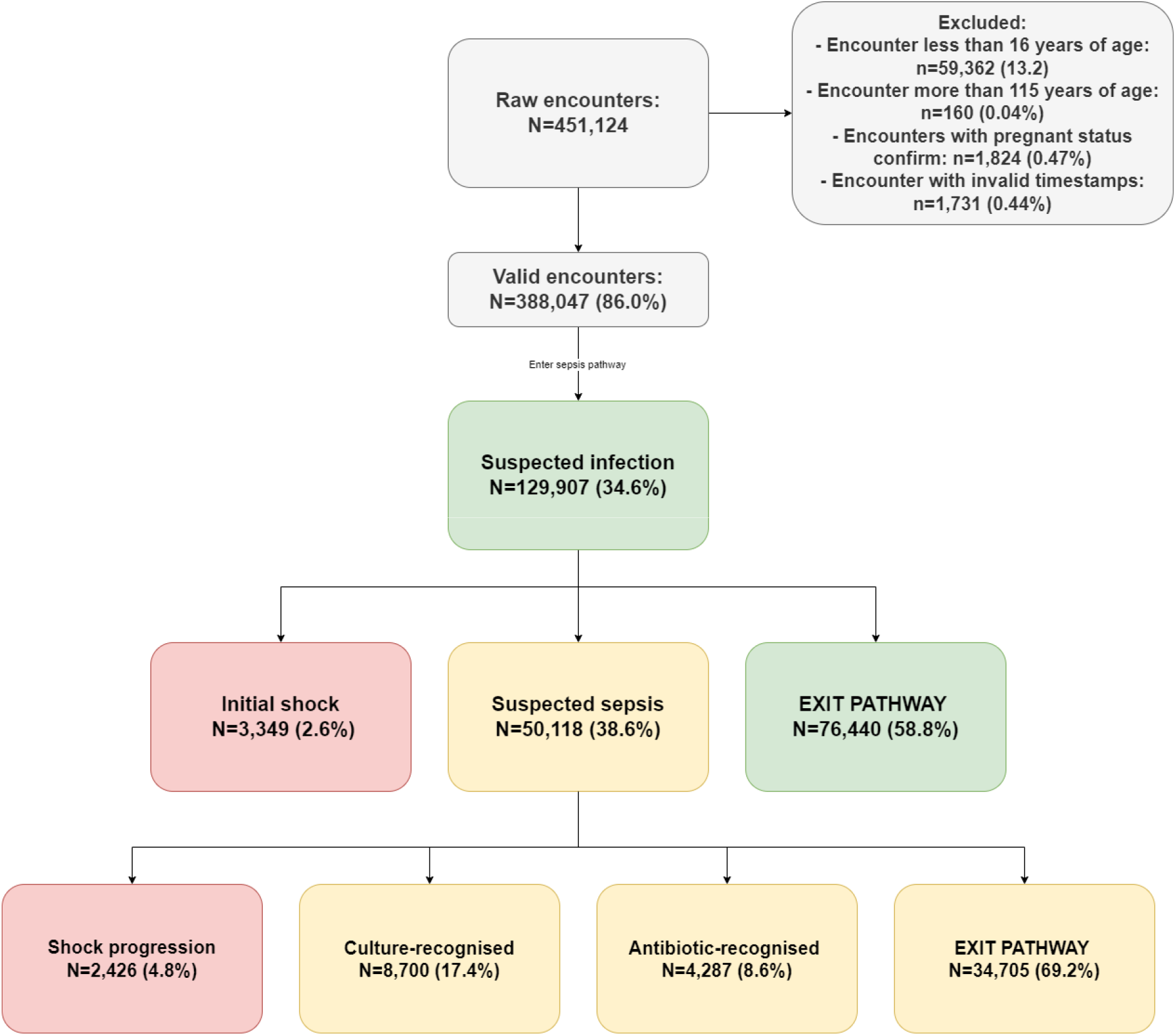
Number and proportion of encounters entering each pathway state The median age was 45 years (IQR 30-65) for all encounters; 197,910 (49.0%) were female. There were 5,967 (1.53%) deaths, and 5,942 (1.53%) encounters involved ICU admission. The median time from ED presentation to first vital measurement was 12 minutes (IQR 8 – 20). After filtering for pathway-relevant events, minimum vital signs for screening were available in 377,189 (97.2%) encounters, and full core vital bundles were available in 273,432 (70.4%) encounters. Essential laboratory tests were available in 265,485 (68.4%) encounters, lactate measurement in 138,798 (35.7%) encounters, blood culture was ordered in 34,055 (8.8%) encounters, IV fluid administration in 113,412 (29.2%) encounters, and antibiotics administration in 74,032 (19.1%) encounters. The median number of events per encounter was 63 (IQR 32–130). These findings indicate that the dataset was sufficiently comprehensive to support encounter-level event reconstruction while also reflecting expected real-world variation in routine documentation.

### Worked example of state-based encounter processing

To illustrate how the architecture operated at the encounter level, Box 1 presents a simplified worked example.

**Box 1.** Worked example of encounter-level processing through the state-based evaluation engine.

A 70-year-old patient presented to the ED with fever and shortness of breath. At triage, an infection-related presenting problem was recorded. Initial observations indicated moderate physiological deterioration, followed by severe abnormality.

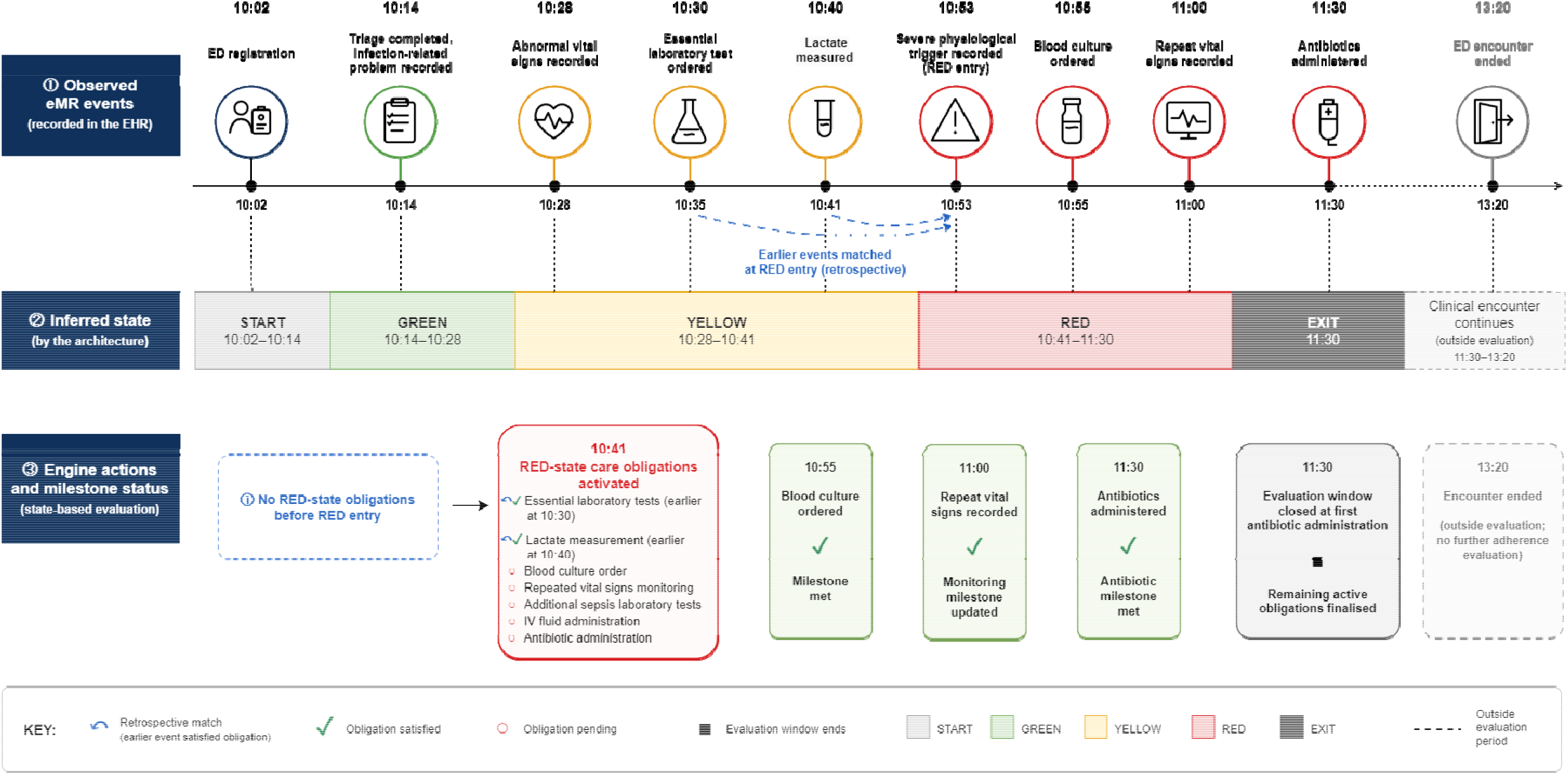

This worked example shows that the architecture did not assign pathway obligations uniformly from presentation. Instead, expected obligations were generated only when the evolving event stream indicated that the patient had entered a clinically relevant state, particularly the RED state.

### Event stream construction and filtering

Across all encounters, a total of 57,631,084 raw eMR events were extracted and transformed into encounter-level event streams. The filtering layer retained 43,951,146 (76.3%) events.

Vital signs accounted for the highest proportion of events, followed by pathology requests and results, medication orders and administrations, and diagnosis-related records (Table 3).

**Table 3.**
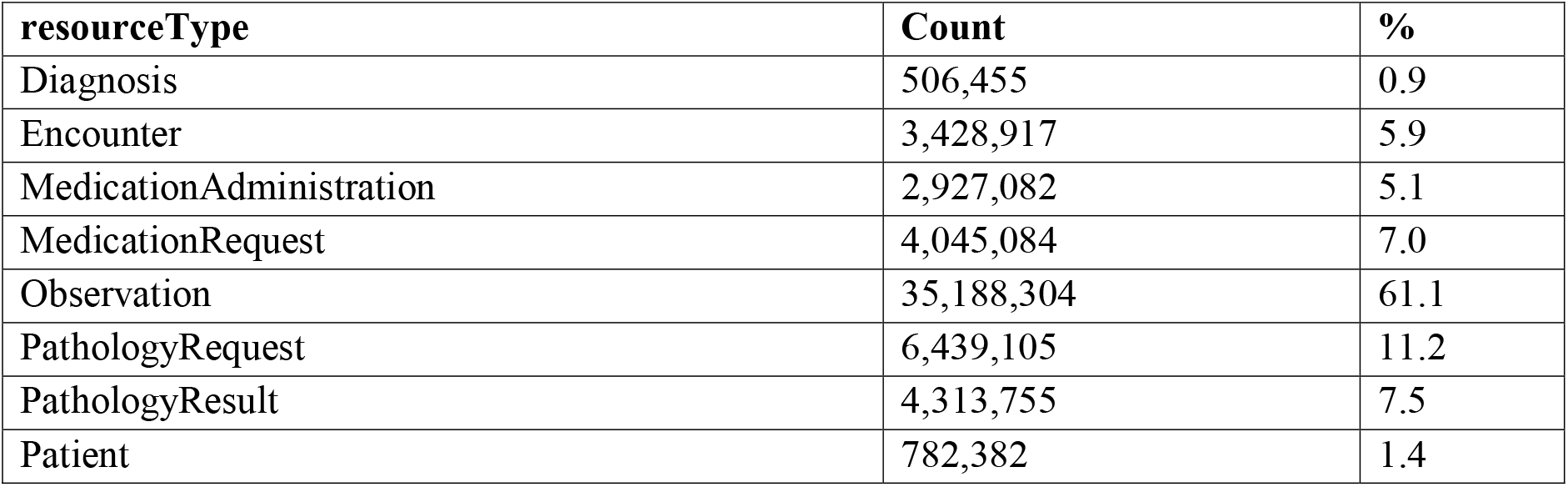
Distribution of retained eMR events by clinical domains.

### Pathway entry and state distribution

Application of state-based pathway logic identified 129,907 (34.6%) encounters that entered the sepsis pathway. Of these, 50,118 (38.6%) progressed to the YELLOW state, indicating increased suspicion of sepsis that requires a senior clinician to confirm, and 3,349 (2.6%) encounters progressed to RED state from GREEN, indicating the initial shock (Figure 2). This demonstrates the architecture’s ability to prioritize higher-acuity states during evaluation.

## Discussion

### Principal findings

In this study, we designed and demonstrated a modular computational architecture for dynamically evaluating adherence to a clinical pathway using routinely collected eMR data. The architecture addresses two key challenges: transforming heterogeneous eMR records into a consistent longitudinal representation and determining which care recommendations apply as a patient’s clinical condition evolves. Application to a sepsis clinical pathway demonstrated that the architecture could evaluating adherence with evolving patient’s condition The principal contribution is a reusable separation between data transformation, pathway-relevant event selection, state determination, temporal adherence evaluation, and downstream deviation analysis. With the rapid change in evidence-based recommendations, this modular design allows pathway criteria to be updated within the evaluation layer without redesigning the entire framework. Additionally, longitudinal event streams can be used to analyze other clinical cases whose best practice standards emphasize the timing and sequence of care recommendations.

Maintaining patient and pathway states was central to dynamic evaluation. These states provide the framework with memory not only of the latest recorded event but also of what has already occurred, making the evaluation process more context-aware. This enabled the architecture to account for care delivered before formal activation of the computational pathway, as well as care recommendations that became applicable only after clinical deterioration or other pathway triggers.

### Relationship to existing computational approaches

The architecture builds on prior work in computer-interpretable guidelines, automated adherence monitoring, process modeling, and conformance checking. These methodologies have demonstrated the feasibility of representing clinical recommendations computationally and comparing them with routinely collected data. [23, 31-33]. However, implementations are often closely coupled with a particular computable guideline format or a predefined process model. Such coupling can limit portability when terminology, clinical criteria, timestamps, or data systems change.

The event-stream transformation layer addresses this issue by converting diverse source records into a uniform chronological format prior to pathway evaluation. This process standardizes records, including observations, investigations, orders, results, and medication administrations, that may use different schemas, local terminologies, and multiple timestamps. This allows evaluation logic to operate independently of the original eMR designs.

The representation of longitudinal clinical events additionally clarifies the relationship between this architecture and process mining approach, which primarily relies on patient event logs for process discovery, conformance checking, and performance analysis [21, 31-33]. While both involve event-based representations of care, many process-mining applications emphasize activity sequences, which are suitable for workflow discovery and control-flow analysis, but may lack sufficient physiological and semantic details needed to capture patient-specific contexts critical for clinical decision-making [31]. A recent scoping review highlights the need for a context-aware, standardized architecture to support scalable, transparent, and real-time adherence monitoring, beyond simple endpoint summaries [34]. The proposed system architecture balances patient-specific temporal trajectories with standardized, computable representations to facilitate clinically relevant conformance evaluation.

### Relationship to interoperability standards

As an emerging interoperability standard, Fast Healthcare Interoperability Resources (FHIR) provides an important standard for exchanging and representing clinical information across systems [35, 36]. Previous work has demonstrated the feasibility of FHIR-based automated guideline-adherence monitoring [24, 37]. However, a FHIR-native representation was not required for the present architecture. The source data was obtained from relational eMR tables rather than a conformant FHIR interface; converting them to FHIR would therefore have added an intermediate representation that would have introduced unnecessary information and implementation complexity for this analytical use case.

Besides, since FHIR is designed for broad interoperability, it permits information to be represented through complex resource hierarchies, optional elements, alternative resources, multiple terminologies, and several timestamps [38-41]. This flexibility supports diverse clinical and technical contexts but does not eliminate the analytical work required before evaluating longitudinal pathways. Data must be structurally normalized into a consistent event schema across different clinical event types, semantically normalized to align local concepts with clinically meaningful categories, and chronologically assembled to reconstruct the sequence of care. Additionally, a consistent event schema is preferred because the adherence engine requires only the clinically meaningful attributes necessary for pathway evaluation, together with separately maintained patient and pathway state. Although FHIR clinical reasoning implementations like Clinical Quality Language [42] or EBM-on-FHIR [43] can describe conditional recommendations and relationships between actions, the execution pathway state required for dynamic pathway evaluation would still need to be implemented within the evaluation framework. The event stream should therefore be viewed as a fit-for-purpose analytical layer rather than an alternative interoperability standard; future implementation could ingest FHIR data and transform it into the same pathway-executable representation.

This distinction is particularly important when comparing the architecture with the work done by Lichtner et al. [24, 43, 44]. Their research program has established an important standards-based approach to computable guideline representation and automated adherence monitoring. Their previous work has shown that a modular approach is better for ease of management and technical implementation [24], with which we share agreement, but this present architecture addresses a different scope of the adherence problem. Lichtner et al.’s monitoring system was designed around individual evidence-based recommendations, typically represented as a condition specifying the target population and an action specifying the recommended intervention. In contrast, the current architecture focuses on progression through an acute clinical pathway in which care recommendations change over time, depending on the patient’s clinical condition and position in the pathway.

### Limitations and future research

Several limitations should be considered. First, the architecture was developed and demonstrated using data from one health service and one acute clinical pathway. Although the components were designed to be separate, portability must be established by applying the event model and evaluation engine to different eMR systems, institutions, and pathways. Second, normalization relied on locally developed clinical mappings. Variation in terminology, units, and documentation practices may require substantial additional mapping at other sites. Third, this architecture relied only on structured eMR data, which may hinder the ability to detect diagnoses or clinicians’ reasoning in unstructured data. Additionally, adherence classifications depended on the completeness and accuracy of eMR documentation. Recorded times may represent ordering, documentation, or system-entry processes. Finally, the study evaluated the technical representation and application of architecture rather than its effect on clinical decision-making, workflow, or patient outcomes. Before using the system to provide real-time feedback or decision support, prospective evaluation is required. For implementation, the system would require ongoing monitoring of data drift, data quality, computational performance, and workflow usability

These limitations can guide future research and development. One improvement is adding more unstructured data, such as free text or imaging reports. Additionally, since the current architecture aligns conceptually with FHIR resource semantics, it provides a pathway to future standardization without requiring a complete rebuild. The present system should thus be viewed as a bridge between legacy local eMR infrastructure and the emerging ecosystem of standardized automated adherence monitoring. The architecture could support future near-real-time implementations, although deployment would require additional work on integration, validation, usability, and governance. While we chose sepsis as the clinical case study, the framework may be adapted to other clinical pathways, since only the relevant pathway logic needs modification, ensuring the system’s generalizability.

## Conclusion

Existing adherence measurement approaches less commonly represent adherence as a dynamic, state-dependent process in which changes in the patient’s clinical condition alter which care recommendations are applicable. In this study, we designed and developed a modular architecture to dynamically evaluate care delivered against best-practice standards using routinely collected eMR data. The architecture transformed heterogeneous eMR data into a consistent longitudinal event stream and used patient and pathway states to evaluate temporally dependent care recommendations, using sepsis as a clinical case. The principal contribution of this study is its modular separation of data transformation and pathway-specific evaluation, which allows pathway logic to be updated without redesigning the entire system, and enables the standardized event streams to be used for other clinical cases. Additionally, maintaining patient and pathway states gives the evaluation framework a record of both the latest events and what has already occurred during an encounter. This enables it to assess adherence to best practices based on the information available at the time of review, providing a more context-aware evaluation that better reflects healthcare delivery. Further multicenter, prospective evaluation is needed to establish portability, validity, and utility for continuous quality monitoring.

## Data Availability

All data produced in the present study are available upon reasonable request to the authors

## Acknowledgments

This research is supported by the Digital Health Cooperative Research Centre Limited (DHCRC). DHCRC is funded under the Australian Commonwealth’s Cooperative Research Centres (CRC) Program. Author MTH holds a higher degree of research scholarship funded through a DHCRC project grant.

## Funding

The funder had no involvement in the study design, data collection, analysis, interpretation, or manuscript writing.

### Declaration of generative AI and AI-assisted technologies in the manuscript preparation process

During the preparation of this work, the author(s) used ChatGPT-4o to support manuscript editing and improve sentence clarity and readability. The author(s) reviewed and edited the output as needed and took full responsibility for the content of the published article.

